# Disgust-Reduction Evaluative Conditioning (DREC) and tDCS in Contamination-Based OCD: A Randomized Controlled Trial

**DOI:** 10.1101/2025.06.04.25328293

**Authors:** Faezeh Raeis Al Mohaddesin, Ali Moghimi, Hamidreza Kobravi, Elahe Yaghoubian, Javad Salehi Fadardi

## Abstract

**Background:** People with contamination-based obsessive-compulsive disorder (C-OCD) experience contamination-related stimuli as more disgusting than normal, which leads to avoidance behavior or washing and cleaning compulsions. This study aimed to investigate whether evaluative conditioning through repetitively pairing a contamination-related stimulus with a pleasant one, and transcranial stimulation over the orbitofrontal cortex (OFC), the brain areas involved in disgust processing, can reduce the intensity of disgust experienced.

**Method:** Forty-eight participants (85% women) with C-OCD were included in a four-armed randomized control trial. They received either sham or active transcranial direct current stimulation (tDCS) over OFC combined with either sham or active disgust-reduction evaluative conditioning (DREC) program for 10 sessions (five days a week). EEG recording, self-report measures, and cognitive tests were performed as pre-, post-, and two-month follow-up assessments.

**Results:** Participants who received active conditioning, active stimulation, or both reported reduced clinical symptoms (*p*≤0.01) and intensity of experienced disgust (*p*≤0.01) after receiving interventions. EEG characteristics changed, especially in theta, alpha, and high-beta frequencies. Inhibitory control improved in groups with active conditioning (*p*≤0.05). There was no change in attentional bias.

**Conclusions:** The current study provided evidence for the effectiveness of DREC and tDCS in modulating disgust feeling and improving C-OCD clinical symptoms. The findings presented serve to highlight the potential of disgust-focused interventions for the treatment of C-OCD. Trial registration: NCT05907369 in the ClinicalTrials.gov.

## 1. Introduction

Disgust dysregulation has a pathological manifestation in contamination-based obsessive-compulsive disorder (C-OCD) that involves an excessive worry about germs and dirtiness, leading to compulsive washing and cleaning behaviors [1,2].

Neuroimaging studies provide evidence supporting the role of the insula and the cortico-striato-thalamo-cortical (CSTC) circuits in both OCD and disgust processing [3]. In OCD, dysfunction of the CSTC leads to increased activity in the orbitofrontal cortex (OFC) [4], a region involved in cognitive appraisal of emotional stimuli [5,6]. The insula, a key region for processing disgust [7], sends information to the OFC [8]. Increased activity and connectivity in the OFC and anterior insula strongly correlate with C-OCD [9]. These neural dysfunctions are often accompanied by significant cognitive impairments, particularly affecting attention and inhibitory control [10,11].

SSRIs and cognitive-behavioral therapy (CBT) focused on exposure-response prevention (ERP), as the first-line treatments for OCD, fail to obtain appropriate responses in approximately half of these patients [12]. Non-invasive brain stimulation (NIBS) methods such as Transcranial direct current stimulation (tDCS) are emerging as research and treatment options for OCD, which has been demonstrated to be significantly effective for symptom improvement [13]. This intervention altered functional brain connectivity and modulates cognitive functions [14,15].

Recent insights have emerged regarding the role of disgust in the pathophysiology and treatment of C-OCD. The evidence supporting that reduced disgust was related to the improvement of C-OCD symptoms [16] has led to the development of a new line of interventions focused on disgust reduction through various methods such as imagery strategies [17] or virtual reality [18].

One approach to altering the emotional value of a stimulus is through evaluative conditioning (EC). EC is a process where pairing a conditioned stimulus (CS) with an unconditioned emotional stimulus (US) can transfer the emotional value of the US to the CS [19]. This mechanism is fundamental in acquiring disgust [20,21]. So, the hypothesis was raised that it might also work in reverse, by pairing a disgusting stimulus with a pleasant one [22]. Importantly, the emotional value established through EC tends to be resistant to extinction [23], which may help explain why exposure therapies are generally less effective in reducing feelings of disgust than fear [24].

Pairing disgusting CS with a pleasant US could reduce its conditioned negative value in individuals with contamination concerns [25]. This method decreased body dissatisfaction [26], which is highly associated with disgust [27]. However, there is limited evidence supporting the effectiveness of this method in clinical samples, and no studies have been conducted specifically on C-OCD.

The current study developed a novel disgust-reduction evaluative conditioning (DREC) program for C-OCD, with the primary objective of attenuating maladaptive disgust responses elicited by symptom-relevant stimuli. We hypothesized that tDCS applied over OFC would show efficacy in disgust modulation, particularly when implemented as an adjunctive neuromodulatory component to the DREC program. This study represents the first randomized controlled trial administering EC combined with tDCS, specifically targeting disgust in patients with C-OCD. Efficacy outcomes were compared across four interventional arms defined by each intervention’s dichotomous administration status (active/sham) (DREC and tDCS) within a 2×2 factorial design framework. We predicted that the most effectiveness would be seen in patients receiving active DREC alongside active tDCS, with either active intervention proving more effective than sham. The hypothesis included that the intensity of disgust experienced and clinical symptoms of C-OCD would be reduced following the interventions (primary outcomes). Given the association between disgust and attentional bias [28], as well as the interference of attentional bias with inhibitory control [29], we investigated whether reducing disgust would improve attentional bias and inhibitory control (secondary outcomes). Quantitative electroencephalography (QEEG) characteristics were also examined (secondary outcome).

## 2. Material and Methods

### 2.1. Study Design

The present study was a randomized controlled trial that administered the DREC program plus tDCS to people with C-OCD in a four-armed design. A comprehensive study design and protocol description can be found in a separate document [30].

### 2.2. Participants

Participants were referred by a psychiatrist or psychotherapist, or self-referred through study ads. All met the DSM-5-TR criteria for OCD with predominant contamination symptoms and provided written informed consent. Inclusion criteria included ages 18-55 and stable medication for the past three months, while exclusion criteria ruled out severe medical or psychological symptoms and any risk for brain stimulation. A total of 48 participants (85% women, M_age_ = 31.1 ± 9.6) completed the trial out of 52 (Fig. 1). Participants received treatment as usual (TAU), including SSRIs and psychotherapy, which needed to remain stable during the trial.

**Fig. 1.**
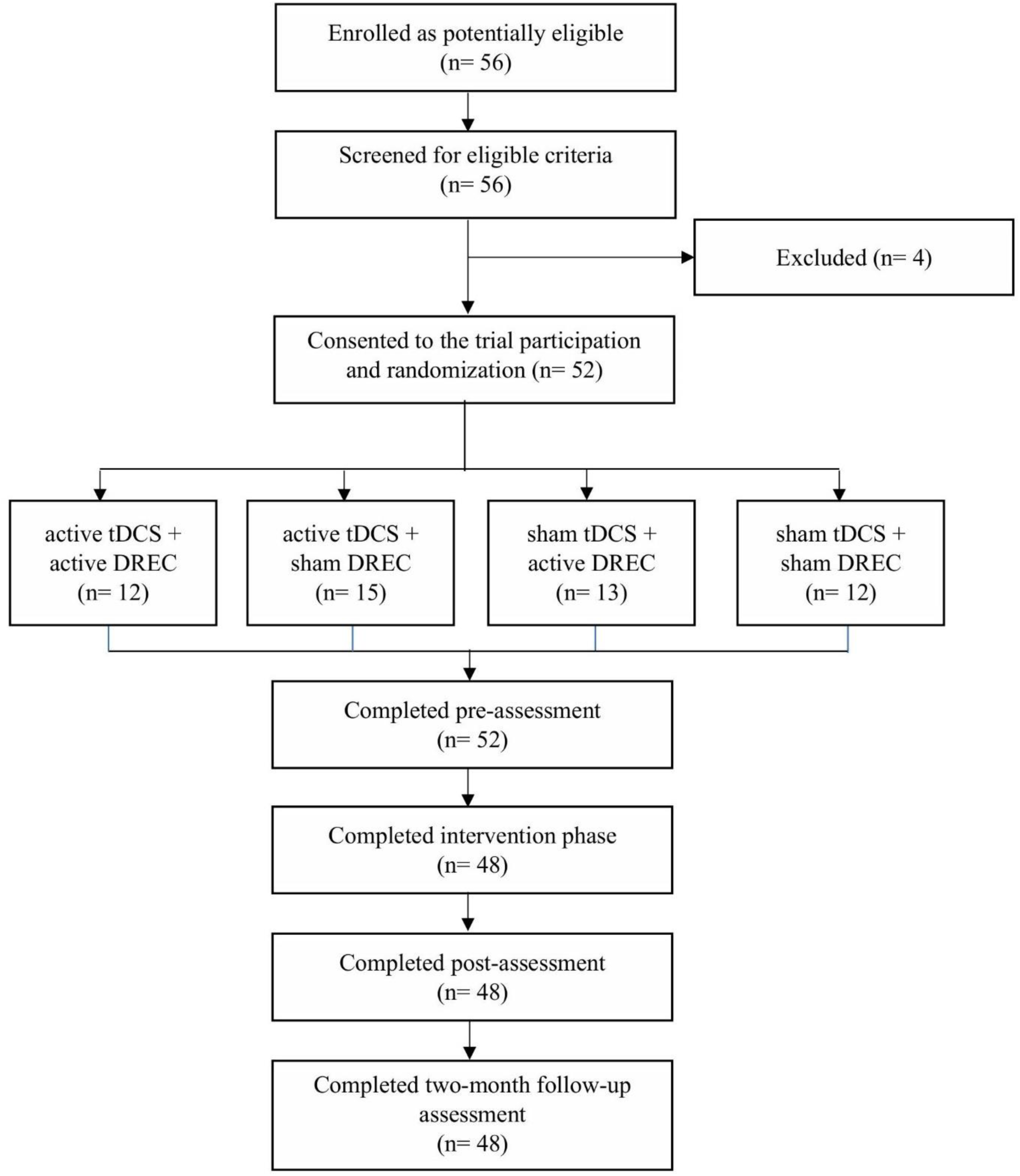
Flow chart.

### 2.3. Interventions

#### 2.3.1. DREC

The DREC program was adapted from Kosinski [26] with several differences: it was computerized rather than a mobile app, included contamination-related pictures as conditioned stimuli (CSs) to target C-OCD symptoms, and provided ten training sessions. The task includes four difficulty levels (Table 1). Thirty contamination-related pictures were paired with 30 pictures as pleasant unconditioned stimuli (pUSs) in the active DREC and with neutral symbols (nUSs) in the sham version, forming 30 specific CS-US pairs, some of which were reviewed in each session. The pictures resulted from a pilot study [see 30]. The pairs were presented in four blocks: the first three blocks displayed a pair list for a limited time, where participants memorized the pairs and matched them in subsequent trials. The fourth block reviewed the pairs without a list. Each session included a total of 240 trials (Fig. 2).

**Fig. 2.**
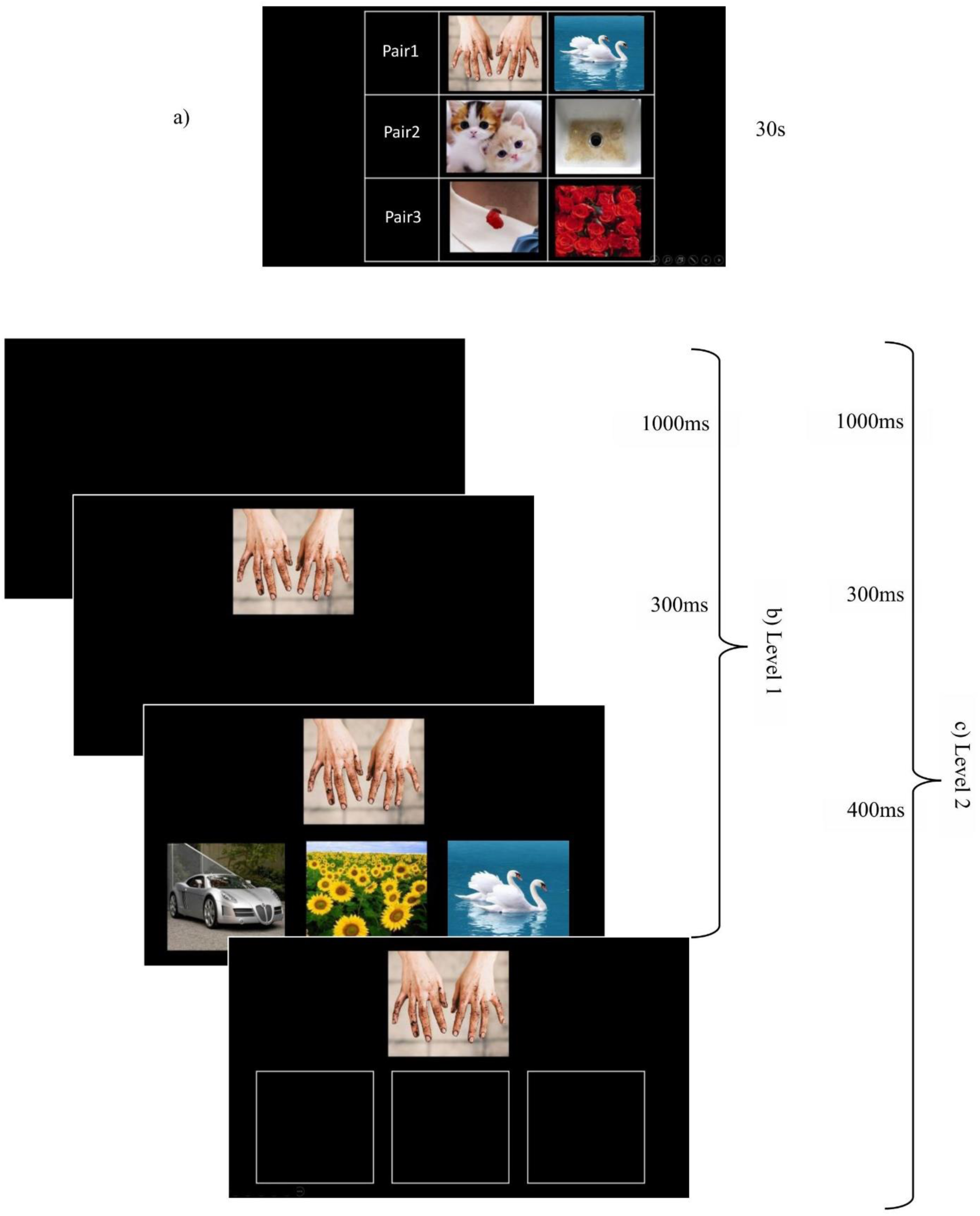
The DREC program: a) an example of a pair list presented at the start of a block for memorization; b) a sample trial for difficulty level 1: a blank screen followed by the CS presented 300 ms before answer options, which remained until a response; c) a trial for difficulty level 2: answer options were replaced with blank frames after 400 ms, remaining until a response.

**Table 1.** The specifications of the DREC program at various difficulty levels.

|  | Level 1 | Level 2 | Level 3 | Level 4 |
| --- | --- | --- | --- | --- |
| Number of Sessions | 3 | 3 | 2 | 2 |
| Number of Pairs in Each Session | 10 | 10 | 15 | 15 |
| Number of Presented Pairs (Blocks1-2-3-4) | 3-3-4-0 | 3-3-4-0 | 5-5-5-0 | 5-5-5-0 |
| Presentation Time for Pair list to be<br>Memorized (Blocks1-2-3-4) | 30s-30s-40s-0 | 30s-30s-40s-0 | 7s-7s-7s-0 | 7s-7s-7s-0 |
| Number of Pairs Repeated for Training<br>(Blocks1-2-3-4) | 3-3-4-10 | 3-3-4-10 | 5-5-5-15 | 5-5-5-15 |
| Number of Repetitions of Each Pair<br>(Blocks1-2-3-4) | 18-18-18-6 | 18-18-18-6 | 12-12-12-4 | 12-12-12-4 |
| Number of Answer Options | 3 | 3 | 5 | 5 |
| Presentation Time for Answer Option | Until the answer | Replaced with blank<br>frames after 400ms | Until the answer | Replaced with blank<br>frames after 400ms |

#### 2.3.2. tDCS

Stimulation was delivered using a battery-driven Activadose (Caputron) tDCS device, with electrodes covered in personalized sponges soaked in 0.9% saline solution. In a bipolar montage, the anode (100 cm²) was placed on the right cerebellum (3 cm below the inion and 1 cm right), and the cathode (35 cm²) was placed on the left OFC (FP1). Active stimulation was set at 2mA for 20 minutes. For sham stimulation, the current was 0.1 mA.

### 2.4. Outcome Measures

#### 2.4.1. Self-report questionnaires

The obsessive-compulsive symptoms were measured using two questionnaires: (1) the Contamination subscale of the Padua Inventory-Washington State University Revision (C-PI-WSUR [31]; and (2) the Yale-Brown Obsessive-Compulsive Scale (Y-BOCS) [32]. The Disgust Rating Scale (DRS) measured the level of disgust experienced from the contamination-related pictures using a visual analog scale (VAS) with ratings from 0 to 10.

Given that depression and anxiety are common comorbid symptoms in OCD patients [33], and disgust propensity (the tendency to experience disgust) correlates with disgust experience in OCD [34], these factors were evaluated at baseline to accurately identify sample characteristics and control for variability in subsequent analyses. The assessments were conducted using the Beck Depression Inventory-II (BDI-II) [35], the Beck Anxiety Inventory (BAI) [36], and the Disgust Scale-Revised (DS-R) [37].

#### 2.4.2. Cognitive measures

Contamination-related and neutral pictures were used to create three computerized cognitive tasks using Psychopy (Psychology software tool, v2021.2.3) [38] to assess attentional bias and inhibitory control as the two prominent cognitive deficits in OCD.

The dot-probe task (DPT) assessed attentional bias toward contamination [39]. It included eight practice trials and 128 main trials, consisting of 96 emotional-neutral pair trials (half congruent, where the probe appeared at the same location as the emotional picture, and half incongruent, where it appeared at the opposite side) and 32 neutral-neutral pair trials. Each trial began with a 500ms fixation cross, followed by a picture pair presented vertically 3 cm apart. After 350 ms, the pictures disappeared, and a probe (the letter F or E) was shown at the location of one of the pictures. Participants pressed the appropriate key as quickly as possible, with an inter-trial interval (ITI) of 1000 ms.

The emotional Go/NoGo task (EGNT) for evaluating proactive inhibition [40] included six practice trials and two experimental blocks, each with six warm-up trials and 96 main trials. A fixation cross appeared for 500 ms, followed by a grey rectangle (in practice and warm-up trials) or a picture (50% contamination-related and 50% neutral) for 250 ms. After the pictures disappeared, a symbol was presented. Participants pressed the response key for symbol $ (Go-trials) and did nothing for symbol # (NoGo-trials). Each subsequent trial began after a 600 ms blank screen (ITI). The presentation order was random, with a 50:50 ratio for Go- and NoGo-trials.

The emotional Stop-signal task (ESST), assessing reactive inhibition [41], began with 16 practice trials, followed by 192 main trials in two blocks. Each trial featured a fixation cross for 500 ms, a picture (contamination-related for half the trials and neutral for the other half) for 100 ms, and a white arrow as the go-stimulus. Participants were instructed to respond quickly to the arrow’s direction (left/right) by pressing the corresponding key (Go-trials; 67%) and to inhibit their response if the arrow turned red (Stop-trials; 33%). In stop-trials, the arrow’s color change served as the stop signal, presented after a fixed stop signal delay (200, 250, 300, or 350 ms). Trials were presented in random order with a 1000 ms ITI.

#### 2.4.3. EEG recording and pre-processing

EEG signals were recorded using a 19-channel cap based on the standard 10-20 EEG electrode system and a Mitsar-202 amplifier, with a 500 Hz sampling rate and a band filter of 0.1 to 30 Hz. Reference electrodes were placed on the earlobes, and all electrode impedances were kept below 10 KΩ. EEG registration lasted three minutes for each of the eyes-closed and eyes-open states.

Data were imported into NeuroGuide Software (Delux v.2.8.1, Applied Neuroscience) and re-referenced to a linked-ear reference. A minimum of 30 seconds of artifact-free signal was selected manually. High-quality selection was verified through split-half and test-retest reliability, with values exceeding 90% and 95%, respectively. Spectral parameters (absolute power, relative power, and peak frequency) from each of the 19 channels and connectivity indices (asymmetry, coherence, and phase lag) from 171 electrode pairs were extracted for delta (1-4 Hz), theta (4-8 Hz), alpha (8-12 Hz), beta (12-25 Hz), and high beta (25-30 Hz) frequencies. Global peak frequency, coherence, and phase lag were calculated as averages from the 19 channels, while regional absolute and relative power were averaged from FP1, FP2, F3, F4, F7, F8 (frontal), Fz, Cz, Pz (midline), and T3, T4, T5, T6 (temporal).

### 2.5. Procedure

Volunteers participated in an initial interview conducted by a clinical psychologist to confirm inclusion criteria, gather demographic information, and obtain written consent. Following baseline assessments, including EEG recording, cognitive measures, and questionnaires, participants were randomly assigned to one of four experimental groups: active conditioning plus active stimulation (aC-aS), sham conditioning plus active stimulation (sC-aS), active conditioning plus sham stimulation (aC-sS), and sham conditioning plus sham stimulation (sC-sS). They completed 10 training sessions (five days a week), each lasting about 25 minutes. The electrodes were placed on the scalp. After one minute of stimulation onset, the DREC program was run. Assessments were repeated after the last training session (post-test) and two months later (follow-up).

### 2.6. Statistical analysis

A sample size of 46 was determined using G*Power 3.1 software for three assessment levels (pre-, post-, and follow-up) with 95% confidence and 80% power, estimating a large effect size with Cohen’s f value of 0.40. Statistical analyses were conducted using SPSS version 29 (IBM Corp., Armonk, NY). To compare primary and secondary outcomes among different intervention groups, a 4 (group: sS-sC, sS-aC, aS-sC, aS-aC) × 3 (time: pre-test, post-test, follow-up) mixed model MANCOVA was employed, treating group as a between-factor and time as a within-factor, with age, BDI, BAI, and DS-R as covariates. The Post hoc Bonferroni test was used for pairwise comparisons, all with a significance level set at *p* < 0.05.

## 3. Results

### 3.1. Descriptive statistics

Table 2 shows clinical characteristics of the groups separately. Each group included 12 members. Among participants, OCD symptom severity was medium for 39.6%, severe for 58.3%, and very severe for 2.1%, according to baseline Y-BOCS. Depression and anxiety symptoms were reported as medium to severe for 62.5% and 77%, respectively. There was no significant difference in baseline clinical characteristics between groups.

**Table 2.**
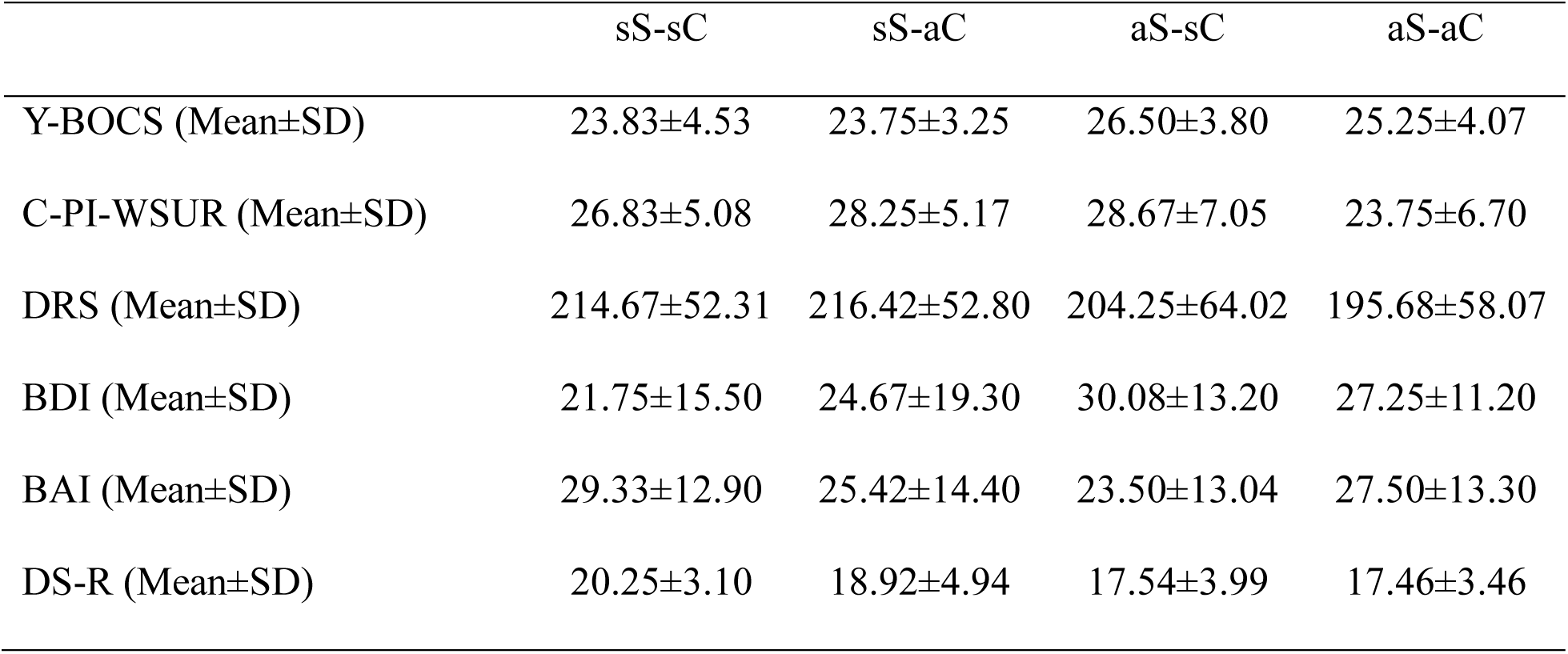
Baseline scores of self-report questionnaires.

### 3.2. Self-report questionnaires

In order to investigate the impact of interventions on clinical C-OCD symptoms and disgust experience, the scores of C-PI-WSUR, Y-BOCS, and DRS were entered into a mixed model MANCOVA as dependent variables. The result showed a significant interaction effect of group×time for the whole model (Pillai’s Trace=0.85, F(18,111)=2.46, *p*=0.002, *η_p_ ^2^*=0.28), and also for C-PI-WSUR (F(6,80)=4.58, *p*<0.001, *η_p_^2^*=0.26), Y-BOCS (F(6,80)=6.27, *p*<0.001, *η_p_^2^*=0.32), and DRS (F(6,80)=3.63, *p*=0.003, *η_p_ ^2^*=0.21), separately. The main effect of the group was significant for Y-BOCS (F(3,40)=3.67, *p*=0.02, *η_p_ ^2^*=0.22).

The post hoc comparisons showed significant differences for all self-report measures across levels for groups with active interventions (sS-aC, aS-sC, and aS-aC), with a reduction for the scores of C-PI-WSUR, Y-BOCS, and DRS in post-test (*p*≤0.01). The reduced score of C-PI-WSUR was sustained until follow-up for these three experimental groups (*p*≤0.01), but Y-BOCS and DRS were preserved just in groups with active conditioning (sS-aC and aS-aC). No significant change was obtained for sS-sC (Fig. 3).

**Fig. 3.**
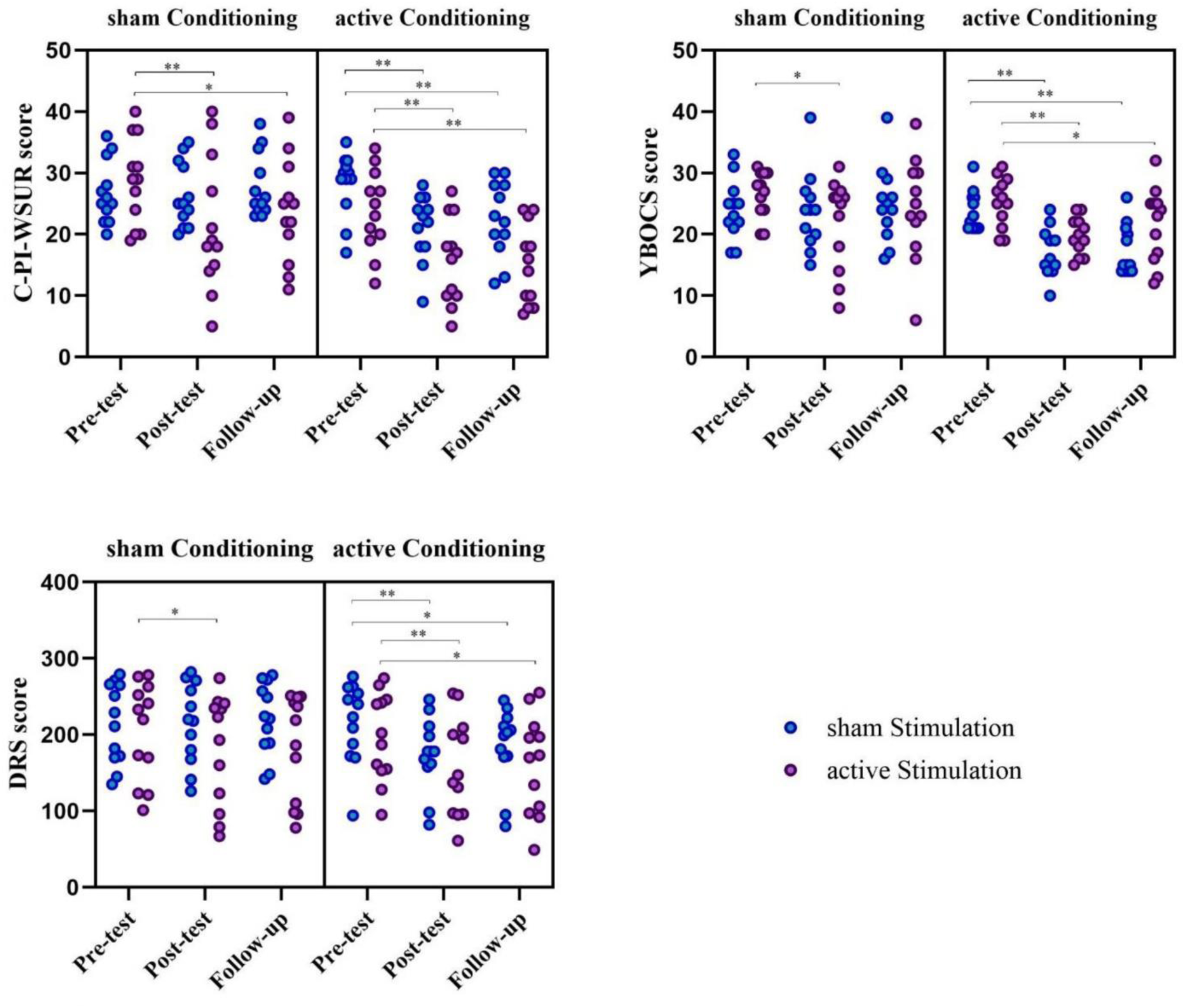
Comparison of self-report measures in different groups across various assessment levels C-PI-WSUR: Contamination subscale of Padua Inventory-Washington State University Revision; Y-BOCS: Yale-Brown Obsessive-Compulsive Scale; DRS; Disgust Rating Scale. \**p*<0.05; ** *p*<0.001

### 3.3. Cognitive measures

Trials with more than 3SD intervals were excluded from the analysis. The Mean reaction time (RT) was calculated from the trials with correct answers. No significant main and interaction effect of group and time was obtained for attentional bias (subtracting the mean RT of congruent trials from incongruent trials) and disengagement (subtracting the mean RT of neutral trials from incongruent trials).

Omission error (no response in Go-trials), commission error (uninhibited response in NoGo-trials), and RT (mean RT in Go-trials) were calculated for disgust trials and neutral trials of EGNT. The interaction of time×DS-R was significant for the model (Pillai’s Trace=0.48, F(12,29)=2.27, *p*=0.03, *η_p_^2^*=0.48) and also for RT in disgust trials (F(1.48,80)=6.36, *p*=0.01, *η_p_^2^*=0.12). Bonferroni post hoc comparison revealed that commission error in disgust trials significantly reduced from pre- (Mean=2.44) to post- (Mean=0.68) in aS-aC group (*p*=0.01). It remained significant until follow-up (Mean=0.26) in this group (*p*=0.005 for pre- to follow-up). Omission error in disgust trials was also statistically significant in the aS-aC group from pre- (Mean=2.09) to post- (Mean=0.97; *p*=0.04).

DVs for ESST included accuracy (% correct in Go-trials), successful inhibition (% correct in Stop-trials), and RT (mean RT in correct Go-trials), separately calculated for disgust and neutral trials. There was significant interaction effect of time×group for successful inhibition (F(6,80)=4.02, *p*=0.001, *η_p_^2^*=0.23) and RT (F(6,80)=3.64, *p*=0.003, *η_p_^2^*=0.22) in neutral trials. Time×BAI showed a significant interaction effect for successful inhibition in neutral trials (F(2,80)=3.63, *p*=0.03, *η_p_^2^*=0.08) and also for RT in neutral (F(2,80)=6.96, *p*=0.002, *η_p_^2^*=0.15) and disgust trials (F(2,80)=4.52, *p*=0.01, *η_p_^2^*=0.10). Time×BDI for RT in neutral trials (F(2,80)=3.56, *p*=0.03, *η_p_^2^*=0.08) and Time×DS-R for RT in neutral (F(2,80)=3.18, *p*=0.047, *η_p_^2^*=0.07) and disgust trials (F(2,80)=5.24, *p*=0.007, *η_p_^2^*=0.12) was also significant. According to the result of post hoc comparison, the sS-aC group showed increased successful inhibition in neutral trials in follow-up (Mean=85.15) compared to pre- (Mean=58.53; *p*<0.001). They also showed increased follow-up RT in neutral trials (Mean=632.06) compared to pre- (Mean=563.15; *p*=0.02).

### 3.4. QEEG characteristics

Power spectral density and connectivity analysis were performed in EC and EO conditions.

The main findings were as follows:

*Absolute power during eyes-closed state*: the absolute powers on midline were changed across time (Pillai’s Trace=0.46, F(10,31)=2.68, *p*=0.02, *η_p_^2^*=0.46), theta in particular (F(1.77,80)=4.39, *p*=0.02, *η_p_^2^*=0.10). The main effect of time is also significant for temporal theta (F(2,80)=3.56, *p*=0.03, *η_p_^2^*=0.08), but the main effect of group for frontal theta (F(3,40)=2.92, *p*=0.04, *η_p_^2^*=0.18). In addition, change in midline high-beta was affected by group×time interaction (F(3.93,80)=3.14, *p*=0.02, *η_p_^2^*=0.19). Frontal high-beta decreased in the aS-aC at follow-up (Mean=1.07) in comparison to post- (Mean=1.83; *p*=0.04); as also in the aS- sC at follow-up (Mean=1.44) in comparison to pre- (Mean=2.73; *p*=0.001). Reduction of midline high-beta was also observed in the aS-aC from post- (Mean=1.40) to follow-up (Mean=0.99; *p*=0.03) and aS-sC from pre- (Mean=2.55) to follow-up (Mean=1.17; *p*<0.001). Temporal beta was statistically reduced in sS-aC from pre- (Mean=13.48) to post- (Mean=9.25; *p*=0.004). See Fig. 4 for an overview.

**Fig. 4.**
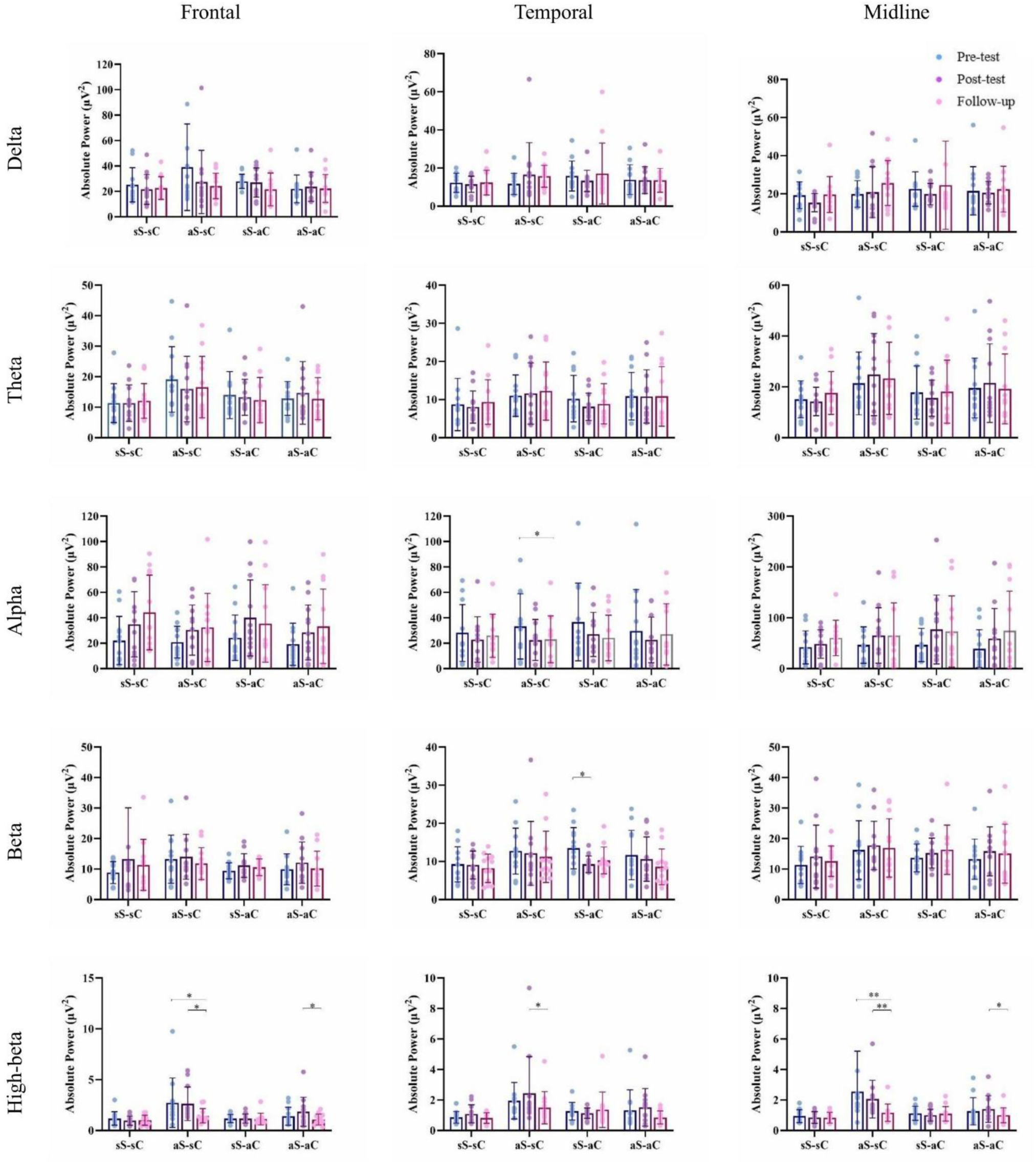
Comparison of the absolute power of the EEG frequency bands, Delta (1-4 Hz), Theta (4-8 Hz), Alpha (8-12 Hz), Beta (12-25 Hz) and High Beta (25-30 Hz) during eyes-closed state. sS-sC: sham stimulation-sham conditioning; aS-sC: active stimulation-sham conditioning; sS-aC: sham stimulation-active conditioning; aS-aC: active stimulation-active conditioning. \**p*<0.05; ** *p*<0.001

*Relative power during eyes-closed state:* midline relative powers were influenced by the main effect of group (Pillai’s Trace=0.57, F(15,114)=1.79, *p*=0.04, *η_p_^2^*=0.19). However, change in midline theta (F(2,80)=4.80, *p*=0.01, *η_p_^2^*=0.11) and temporal theta (F(2,80)=4.46, *p*=0.02, *η_p_^2^*=0.10) were significant across time, and midline high-beta across group (F(3,40)=4.01, *p*=0.01, *η_p_^2^*=0.23). Time×BAI interaction for frontal delta (F(2,80)=3.55, *p*=0.03, *η_p_^2^*=0.08) and midline delta (F(1.59,80)=4.15, *p*=0.03, *η_p_^2^*=0.09), and time×DS-R interaction for frontal theta (F(2,80)=3.83, *p*=0.03, *η_p_^2^*=0.09) were also significant. Further post hoc comparison revealed decrease in frontal and midline theta from pre- (Mean=19.70, 22,16) to post- (Mean=14.01, 13.79) in aS-aC (*p*=0.007, 0.005), decrease in frontal and temporal high-beta from post- (Mean=2.93, 3.74) to follow-up (Mean=1.80, 2.45; *p*=0.001, 0.02) and midline high-beta from pre- to follow-up (Mean=2.44, 1.20, *p*=0.04), in aS-sC.

*Absolute power during eyes-open state:* a significant time×group interaction effect was obtained for midline beta (F(6,80)=3.40, *p*=0.005, *η_p_^2^*=0.20) and high-beta (F(6,80)=2.57, *p*=0.03, *η_p_^2^*=0.16) and also for temporal beta (F(4.94,80)=2.49, *p*=0.04, *η_p_^2^*=0.16) and high-beta (F(2,80)=2.38, *p*=0.04, *η_p_^2^*=0.15). Time for temporal theta was significant (F(2,80)=3.29, *p*=0.04, *η_p_^2^*=0.08), but the effect of time for temporal high-beta was dependent on DS-R (F(2,80)=7.47, *p*=0.001, *η_p_^2^*=0.16). Based on post hoc analysis, midline beta increased from pre- to post- (Mean=9.85, 12.50) in aS-aC (*p*<0.001) and from post- (Mean=8.69) to follow-up (Mean=10.46) in sS-sC (*p*=0.04), but midline high-beta decreased from pre- (Mean=2.36) to follow-up (Mean=1.34) in aS-sC (*p*<0.001). The aS-sC group also showed increased temporal theta from pre- (Mean=6.45) to post- (Mean=9.00, *p*=0.01) and follow-up (Mean=9.36, *p*=0.008) and decreased temporal high-beta from pre- (Mean=1.87) to follow-up (Mean=1.21, *p*=0.02). An acute increase in temporal alpha was observed in aS-aC from pre- (Mean=12.54) to post- (Mean=16.10, *p*=0.03). The temporal alpha increase was also significant in sS-sC, but from post- (Mean=11.15) to follow-up (Mean=17.38, *p*=0.04). See Fig. 5 for an overview.

**Fig. 5.**
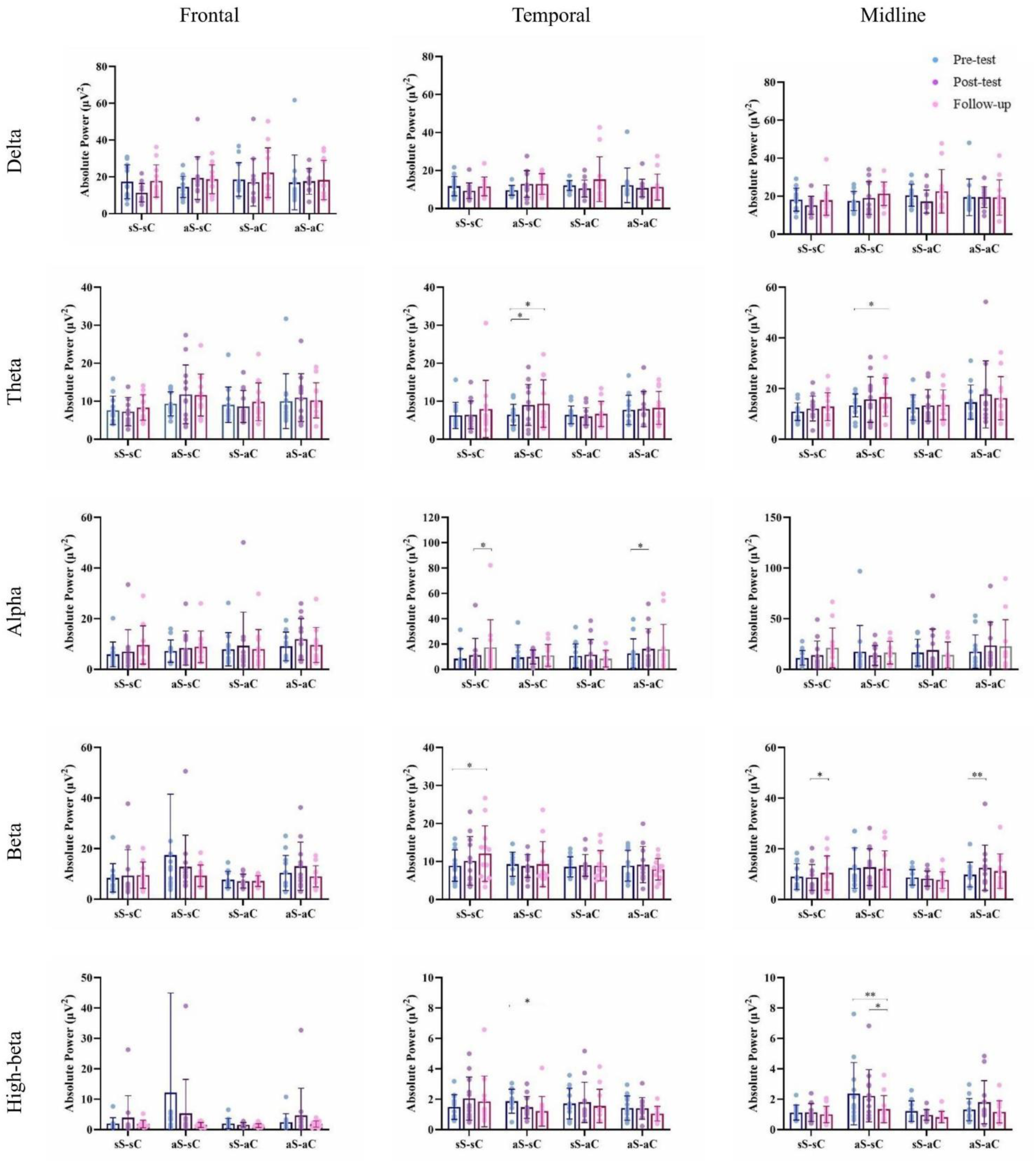
Comparison of the absolute power of the EEG frequency bands, Delta (1-4 Hz), Theta (4-8 Hz), Alpha (8-12 Hz), Beta (12-25 Hz) and High Beta (25-30 Hz) during eyes-open state. sS-sC: sham stimulation-sham conditioning; aS-sC: active stimulation-sham conditioning; sS-aC: sham stimulation-active conditioning; aS-aC: active stimulation-active conditioning. \**p*<0.05; ** *p*<0.001

*Relative power during eyes-open state:* Time×group interaction was at a significant border for frontal theta (F(6,80)=2.21, *p*=0.05, *η_p_^2^*=0.14), and was statistically acceptable for temporal delta (F(6,80)=2.28, *p*=0.04, *η_p_^2^*=0.15), theta (F(6,80)=5.61, *p*<0.001, *η_p_^2^*=0.30), beta (F(6,80)=2.33, *p*=0.04, *η_p_^2^*=0.15), and high-beta (F(5.04,80)=3.55, *p*=0.02, *η_p_^2^*=0.18). The time effect for temporal theta was significant in interaction by BDI (F(2,80)=5.08, *p*=0.008, *η_p_^2^*=0.11) and also by BAI (F(2,80)=3.27, *p*=0.04, *η_p_^2^*=0.07), and time×DS-R interaction effect was obtained for temporal high-beta (F(2,80)=4.98, *p*=0.009, *η_p_^2^*=0.11). According to further analysis, sS-aC showed post- (Mean=35.50) to follow-up (Mean=43.02) increase in frontal delta (*p*=0.01); From pre- to follow-up, aS-sC showed increase in frontal theta (Mean=18.26, 22.15; *p*=0.02), and temporal theta (Mean=16.40, 20.03; *p*<0.001), and decrease in frontal beta (Mean=22.10, 18.42; *p*=0.01), frontal high-beta (Mean=7.07, 3.12; *p*=0.008), temporal high-beta (Mean=4.66, 2.88; *p*<0.001), and midline high-beta (Mean=3.62, 2.09; *p*=0.001). This group also showed midline beta decrease from post- (Mean=19.43) to follow-up (Mean=16.98; *p*=0.006). Opposed to aS-sC, temporal theta decrease was observed in sS-sC from pre- (Mean=16.26) to follow-up (Mean=14.51; *p*=0.005). Additionally, midline alpha increased from post- (Mean=22.75) to follow-up (Mean=27.61; *p*=0.02), and temporal delta decreased from pre- (Mean=31.72) to follow-up (Mean=26.29; *p*=0.03) in sS-sC.

*Peak Frequency:* A significant interaction effect of time×group was observed on peak frequency during eyes-closed state (Pillai’s Trace=1.00, F(30,99)=1.65, *p*=0.03, *η_p_^2^*=0.33) particularly for theta (F(6,80)=3.28, *p*=0.006, *η_p_^2^*=0.20), and alpha (F(4.38,80)=3.08, *p*=0.02, *η_p_^2^*=0.19). The aS-sC group showed a pre- (Mean=2.22) to follow-up (Mean=2.09) decrease in peak frequency of delta (*p*=0.02) and a pre- (Mean=9.64) to post- (Mean=9.52) decrease in peak frequency of alpha (*p*=0.03) in the eyes-closed state. Moreover, high-beta peak frequency during the eyes-closed state decreased from post- (Mean=27.07) to follow-up (Mean=27.00) in the sS- aC group (*p*=0.04), and eyes-open beta (Mean=17.50, 16.79; *p<*0.001) and high-beta peak (Mean=27.08, 26.98; *p*=0.02) decreased from pre- to follow-up in the aS-sC group.

*Connectivity:* The main effect of group showed significance for eyes-open temporal asymmetry, considering all band frequencies as DVs (Pillai’s Trace=0.63, F(15,114)=2.04, *p*=0.02, *η_p_^2^*=0.21), and the main effect of time was also nearly significant for EO frontal alpha asymmetry (F(1.61,80)=3.46, *p*=0.047, *η_p_^2^*=0.08). Frontal high-beta asymmetry during the eyes-closed state was changed in the aS-aC group with the left dominance in pre-test (Mean=20.90) but a slight right dominance in follow-up (Mean=-3.43; *p*=0.04).

Investigating eyes-open coherence revealed a significant main effect of time for beta (F(1.75,80)=4.02, *p*=0.03, *η_p_^2^*=0.09), and high-beta (F(2,80)=5.41, *p*=0.006, *η_p_^2^*=0.12), and a main effect of group for theta (F(3,40)=2.88, *p*=0.048, *η_p_^2^*=0.18). The aS-sC group (Mean=36.18) showed less post-theta coherence during the eyes-open state than the sS-sC group (Mean=43.91; *p*=0.04). No significant result was obtained for coherence and phase lag during the eyes-closed state, and phase lag coherence during the eyes-open state.

## 4. Discussion

The crucial role of disgust in the development and treatment of OCD is clear, but how to modulate it is yet a debate. This was the first study to investigate the effectiveness of EC in the framework of the DREC program and tDCSC-OFC on disgust experienced in C-OCD samples besides their clinical symptoms, EEG characteristics, and cognitive functions.

Our findings indicate that DREC and tDCS reduced the intensity of disgust experienced and clinical symptoms. Cathodal tDCS over the OFC is a well-established brain stimulation method for treating OCD [42,43], and this study demonstrated its effectiveness in reducing disgust experiences as well. Given the OFC’s significant role in value representation [44], decreased cortical excitability through cathodal stimulation may reduce overestimated disgust values by modulating the hyperactive OFC and its associated cortical and subcortical areas.

The disgusting value was diminished after tDCS, but it was not lasting two months later. In contrast, the reduced disgust experience remained evident at follow-up in the groups that received active DREC. As hypothesized, repetitive pairing of contamination-related pictures with a pleasant US helped modulate their negative value. According to the declarative memory model [45], pairing CS-US creates a memory trace that can be consciously retrieved during later evaluations. Therefore, the more consolidated the memory, the more stable the EC effect. In the present study, repeating EC training across sessions aimed to create a strong memory trace. Additionally, the training program utilized active recall to enhance new associative learning [46].

Additionally, the DREC program, both with and without tDCS, effectively decreased participants’ scores on OCD questionnaires. The main assumption of the study was that reducing the heightened feeling of disgust would effectively improve compulsive behaviors and unnecessary avoidance. Since disgust is particularly resistant to extinction, especially regarding high contamination concerns [47], this approach was deemed more beneficial [48]. This finding contrasts with studies that indicate evaluative conditioning does not exceed mere exposure by only changing expectancy without affecting the disgusting value of conditioned stimuli [49] or avoidance behavior [50]. However, it aligns with the findings demonstrating that evaluative conditioning employing positive USs effectively reduced conditioned disgust in individuals with both high and low contamination concerns [25].

Neurophysiological changes account for improving C-OCD symptoms after tDCS [51], whereas generalization may better explain improvements following DREC. According to generalization theory, the acquired value through EC can extend to other stimuli within the same category as CS [52,53]. This generalization primarily occurs when a CS has a prominent feature (e.g., contamination) that gets selective attention [54]. It appeared that the EC effect extended to real-life situations that trigger OCD symptoms. Interestingly, two participants declared they would automatically recall pleasant pictures when exposed to contamination-related situations.

Although DREC and tDCS effectively reduced the value of disgusting stimuli, the attentional bias measured by DPT did not change. Typically. Individuals with contamination concerns exhibit an attentional bias toward disgust-related stimuli [55], but neither DREC nor tDCS directly impacted this bias. One possible explanation is that the pictures were still perceived as disgusting compared to neutral pictures. It is essential to understand that a disgusting contamination-related stimulus never acquires a positive valence. Furthermore, EC may not be even strong enough to make them neutral, especially in individuals with OCD who have high disgust sensitivity [56]. The other explanation concerns fear. While participants rated pictures based on disgust, we do not have information about the intensity of experienced fear. The fear that is provoked could lead to an attentional bias, particularly since it is often the most familiar emotion experienced in individuals with OCD, and exposure is the most effective method for reducing it [57].

The improvement in inhibitory control was limited. Proactive inhibition (EGNT) toward disgusting stimuli improved following DREC plus tDCS, and reactive inhibition (ESST) toward neutral stimuli following active DREC. As expected, the larger effect was obtained from the combination of interventions. There was no change following tDCS alone, and the only outcome from DREC was at the expense of slower reaction times.

Considering the improvement of symptoms after interventions, despite a lack of or slight change in cognitive performance, the assumption that comes to mind is that cognitive functions and clinical symptoms may not align as expected, contrary to our initial hypotheses. We observed a reduction in clinical symptoms without changes in attentional bias; the others [58] found no change in avoidance behavior despite a reduced attentional bias. The result aligns with the evidence indicating a weak association between OCD symptom severity and cognitive functions [10] or persistent cognitive deficits despite symptom improvement, which supports a trait-like approach to the cognitive deficits associated with OCD [59]. The final explanation is that participants’ performance on cognitive measures initially did not differ from normal, as some studies found no significant difference in cognitive performance between individuals with OCD and healthy control participants [60]. However, we could not test this in our study due to the lack of a healthy group.

The QEEG data revealed two main findings: 1) notable changes in theta and high-beta frequency bands, and 2) more extensive changes at the two-month follow-up compared to the post-test. Theta relative power decreased in both frontal and midline areas during the eye-closed state after tDCS plus DREC. This decrease could indicate successful treatment since increased power in slow frequency bands is a potential biomarker [61]. Other studies have also reported decreased slow frequency power after tDCS [62] and TMS [63]. Research findings regarding alpha and beta activity in OCD are inconsistent [64]. However, temporal alpha increased after tDCS plus DREC, alpha peak frequency decreased after tDCS, and temporal beta decreased after DREC.

At follow-up, we observed a decrease in the absolute power of high-beta in the frontal and midline regions for groups receiving active tDCS, whether combined with active or sham DREC. Additionally, high-beta relative power decreased in the aS-sC group. More findings revealed a reduction in delta, beta, and high-beta peak frequency in the aS-sC group, as well as a decrease in high-beta peak frequency in the sS-aC group. The significant results observed at follow-up, which were not seen at the post-test, suggest that changes in EEG characteristics may become more evident over time.

The current study has several limitations, including the relatively small sample size with almost ethnic homogeneity. Most of the participants were female. Pharmacotherapy or psychotherapy during the trial may influence the effect of interventions. To improve generalizability, future research is needed to replicate these findings, particularly in larger samples, unmedicated patient groups from different ethnicities and cultures. Determining the optimal dose of intervention sessions is crucial for effectiveness. We suggest combining EC with other tDCS montages and another stimulation method, such as repetitive transcranial magnetic stimulation (rTMS), to enhance our understanding further. Employing other cognitive tasks may yield more information about cognitive changes. Designing alternative EC programs (beyond DREC), potentially as a mobile app, warrants exploration. Individualized EC material holds promise for significantly improving results. Future studies could also explore these interventions for other OCD subtypes or disorders where disgust plays a significant role.

## 5. Conclusion

This study provided a new computer training program using EC to reduce disgust. It aimed to focus on the clinical implications of disgust in OCD treatment and demonstrated that EC can be an appropriate method for modulating disgust in the C-OCD population. EC as a complementary treatment, aside from other methods such as brain stimulation, may create hope to optimize treatments for people suffering from such a disabling disorder.

## Statement of ethics

The study was approved by the ethics committee of Ferdowsi University of Mashhad (IR.UM.REC.1400.350).

## Data Availability

Data is publicly available at Mendeley Data.

https://doi.org/10.6084/M9.FIGSHARE.C.6947452

## Acknowledgements

We would like to thank all the members of the Cognitive Science Lab (FUM) and Aren Center for their invaluable help in data acquisition. We would like to thank Fatemeh Asadollahzadeh for her help in preprocessig EEG data.

## Data sharing

Data is publicly available at Mendeley Data (doi:10.6084/M9.FIGSHARE.C.6947452).

